# SARS-CoV-2 transmission and control in a hospital setting: an individual-based modelling study

**DOI:** 10.1101/2020.08.22.20179929

**Authors:** Qimin Huang, Anirban Mondal, Xiaobing Jiang, Mary Ann Horn, Fei Fan, Peng Fu, Xuan Wang, Hongyang Zhao, Martial Ndeffo-Mbah, David Gurarie

**Affiliations:** Department of Mathematics, Applied Mathematics and Statistics, Case Western Reserve University, Cleveland, OH, USA, 44106.; Department of Neurosurgery, Union Hospital, Tongji Medical College, Huazhong University of Science and Technology, Wuhan, China, 430022.; Department of Veterinary and Integrative Biosciences, College of Veterinary and Biomedical Sciences, Texas A&M University, College Station, TX, USA, 77840; School of Public Health, Texas A&M University, College Station, TX, USA, 77840.; Center for Global Health and Diseases, School of Medicine, Case Western Reserve University, Cleveland, OH 44106, USA

**Keywords:** COVID-19, non-pharmaceutical interventions, hospital setting, individual-based model

## Abstract

**Background:** Development of strategies for mitigating the severity of COVID-19 is now a top global public health priority. We sought to assess strategies for mitigating the COVID-19 outbreak in a hospital setting via the use of non-pharmaceutical interventions such as social distancing, self-isolation, tracing and quarantine, wearing facial masks/ personal protective equipment.

**Methods:** We developed an individual-based model for COVID-19 transmission among healthcare workers in a hospital setting. We calibrated the model using data of a COVID-19 outbreak in a hospital unit in Wuhan in a Bayesian framework. The calibrated model was used to simulate different intervention scenarios and estimate the impact of different interventions on outbreak size and workday loss.

**Results:** We estimated that work-related stress increases susceptibility to COVID-19 infection among healthcare workers by 52% (90% Credible Interval (CrI): 16.4% – 93.0%). The use of high efficacy facial masks was shown to be able to reduce infection cases and workday loss by 80% (90% CrI: 73.1% – 85.7%) and 87% (CrI: 80.0% – 92.5%), respectively. The use of social distancing alone, through reduced contacts between healthcare workers, had a marginal impact on the outbreak. A strict quarantine policy with the isolation of symptomatic cases and a high fraction of pre-symptomatic/ asymptomatic cases (via contact tracing or high test rate), could only prolong outbreak duration with minimal impact on the outbreak size. Our results indicated that a quarantine policy should be coupled with other interventions to achieve its effect. The effectiveness of all these interventions was shown to increase with their early implementation.

**Conclusions:** Our analysis shows that a COVID-19 outbreak in a hospital’s non-COVID-19 unit can be controlled or mitigated by the use of existing non-pharmaceutical measures.

## Background

The world is in the midst of an unprecedented coronavirus outbreak caused by a novel virus recently named COVID-19 by the World Health Organization (WHO). The outbreak which started in the city of Wuhan, Hubei Province, China, in early December 2019 spread to many countries around the world before being declared a pandemic by the WHO on March 11th, 2020.

Developing strategies for mitigating the severity of COVID-19 is now a top global health priority. The range of containment strategies employed in different countries and regions varies from shelter-in-place orders, the shutdown of public events, travel ban [1], and visitor quarantine, to intermediate steps that involve partial closures (e.g. schools [2], workplaces, sporting, and cultural events) [3]. While such drastic steps can reduce infection spread, they exact a heavy toll on society and human well-being. At present the only available means of containing COVID-19 spread is via the use of non-pharmaceutical interventions [4, 5] such as social distancing, self-isolation [6], tracing and quarantine [6, 7], wearing facial masks/ personal protective equipment (PPE) [8, 9].

Mathematical models of disease transmission are powerful tools for exploring this complex landscape of intervention strategies and quantifying the potential benefits of different options [10–13]. Traditional approaches in epidemiological modeling use compartmental models [14–16], which assume a uniform population and simple mixing patterns with steady contact rates. Such models can give qualitative answers for large-scale populations at best [17, 18], however, are not suitable to account for the complexity and specifics of COVID-19 in local communities and small populations (e.g., hospital, workplace, school). Such settings are characterized by heterogeneous populations, multiple disease pathways, and complex social interactions.

Our focus here is COVID-19 transmission in a hospital setting, where healthcare workers (HCWs) are at high risk to acquire infection through interactions with fellow HCW and with patients [19–22]. We developed a novel individual-based model (IBM) for COVID-19 transmission among HCWs, and applied it to explore the efficacy of different control/ mitigation strategies via non-pharmaceutical interventions. IBMs have been used extensively to model pathogens spread on different scales, from global pandemics [23–25], to local social networks [26]. On the disease side, our IBM features distinct infective stages and transitions, observed in Covid-19, with some hosts recovering without any symptoms, while others undergoing mild or severe infection pathways. On the social side, we take into account individual behavior, including mixing patterns among HCW, their use of facemasks and PPE, and HCW-patient interactions. All of these factors play an important role in COVID-19 transmission.

The IBM model was calibrated in a Bayesian framework using empirical data from a non-COVID hospital unit. We used our calibrated model to simulate different intervention scenarios, including adaptive behavior (social distancing in the workplace, individual protection, isolation of infected individuals). In each case, we assessed the effect of interventions on outbreak outcomes: outbreak size, outbreak duration, and workday loss.

## Methods

### Individual-based modelling methodology

In our model, an individual can undergo a sequence of infection stages, classified as susceptible (*S*), pre-symptomatic/ asymptomatic (*E*), two symptomatic stages *I*_1_ (upper respiratory stage) followed by *I*_2_ (advanced infection stage, lungs et al.), and recovered/immune state (*R*) (see Figure 1). These states differ by their infectivity levels and stage duration. Unlike most other viral diseases, pre-symptomatic/asymptomatic COVID-19 hosts (*E*-stage) are known to transmit pathogens [27–29]. So we assign positive infectivity levels (*b*_0_, *b*_1_, *b*_2_) to all three stages(*E*, *I*_1_, *I*_2_).

**Figure 1:**
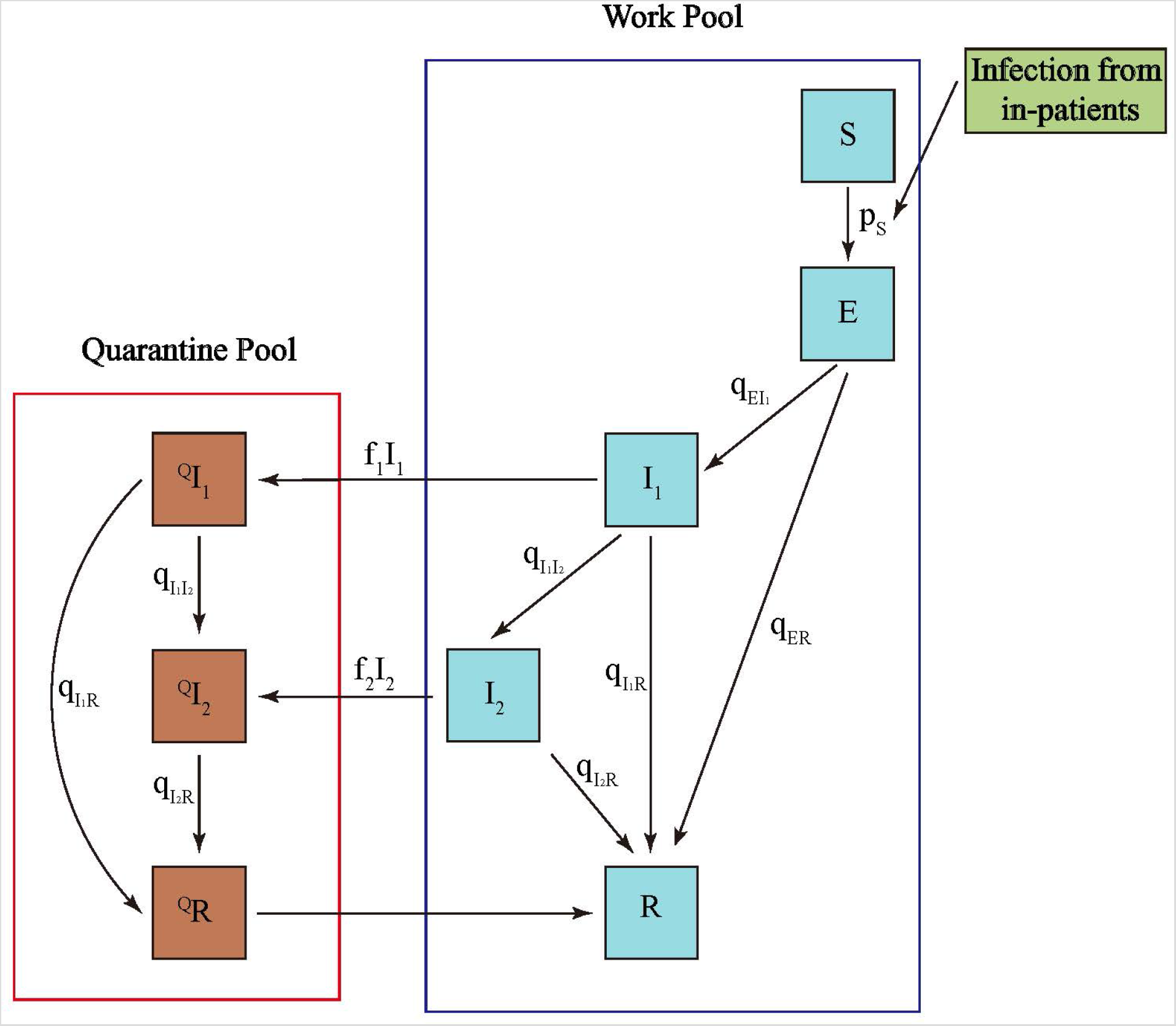
State transitions in the individual-based model. The standard SEIR scheme is used to describe host states: (*S*)- susceptible, (*E*)- latent (pre-symptomatic/ asymptomatic but infectious), *I*_1_- first symptomatic (upper respiratory infection), *I*_2_- second stage (advanced lung infection), (*R*)- recovered/immune. Hosts can undergo three different pathways: asymptomatic(*E* → *R*); mild symptomatic(*E* → *I*_1_ → *R*); severe symptomatic(*E* → *I*_1_ → *I*_2_ → *R*). Depending on the screening procedure, fractions of (*I*_1_, *I*_2_) are sent to quarantine, and released to the workpool upon recovery. Infected patients are treated as an external source

We modelled social mixing patterns by assuming that HCWs and ward patients interact on a daily basis via aggregating in random groups of HCWs, and via patient visitation by HCWs (See SI in details). The net outcome is a contact pool for each HCW-host, which varies randomly on a daily basis. Each contact of a susceptible individual with infectious individuals (HCWs or patients) can lead to infection (transition (*S* → *E*), with a probability that depends on infectivity levels of the contact pool and the host susceptibility, *a*(*a* = 0 - fully protected, *a* = 1 - fully susceptible). The latter depends on host health /immune status, individual behavior e.g. use of facial masks, and environmental conditions. For instance, HCWs are supposed to use additional protection when contacting patients. Then a probability of ‘surviving’ a single infective contact *b*_i_ for an *S*-host of susceptibility *a*, is given by 1 - a *b*_i_. Combining all infective contacts of a given *S-*host, we get the probability of infection (*S* → *E*), 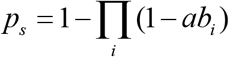.

We divided all HCWs staff into susceptibility strata based on the hospital data [30]: (i) normal pool, 60% of HCWs, have baseline susceptibility value, *a_N_* = 0.5; (ii) high-risk (stressed) pool, 40% of HCWs, with susceptibility level, 0.5 < *a_S_* < 1 (to be calibrated).

Two points of our setup require some clarification: (i) the proposed form of social mixing in random clusters extends the conventional ‘social network’ transmission pathways (see Figures. S1–S2); (ii) an infective ‘social contact’ in our context means an event of sufficient duration and proximity, to allow transmission of pathogens from infected to susceptible host [31].

There is much uncertainty on disease progression of infective stages. Here we assume infected *E-*hosts can undergo three different pathways: (A) asymptomatic *(*E → R*); (*M*) mild symptomatic (*E → I_1_ → R*); (S) severe symptomatic (*E → I_1_ → I_2_ → R*), with population fractions (*v_A_;v_M_;v_S_*). In all cases, pre-symptomatic/ asymptomatic pool (*E*) can carry and transmit the virus, along with (*I_1_ I_2_*). Each infective stage (*E,l_x_,l_2_*) has associated (mean) duration, *L_E_; L_1_; L_2_*. The probability of state transition (*E → I_1_; I_1_ → I_2_*, etc.) depends on the time, *d*, spent in a given disease state, relative to the mean stage duration, L. Specifically, we use a Bernoulli distribution *B*(*p*) of parameter, *p* = *Φ* (*d/L*), with sigmoid (0 < *Φ* (*x*) < 1*) of half-value, *x* = 1/2.

During an outbreak, the HCWs expressing symptoms are tested, and certain fractions (*f_1_; f_2_*) of (*I_1_; I_2_*) are put in isolation, where they undergo their specific disease pathways, but do not mix and transmit the pathogen. Two different types of diagnostic tests were used in the hospital, PCR for light symptoms, and lung-scan for more severe conditions [30, 32]. Thus our assumed quarantine fractions (0 < *f_i_* < 1) account for limited test sensitivity, and a possible overlap of ‘COVID-like’ symptoms, expressed by non-COVID hosts. The recovered HCWs return to the work pool (see Figure 1).

The model simulations were run on a daily basis and implemented in the Wolfram Mathematica platform. The key inputs in the model include: 1) population makeup in terms of asymptomatic, mild, and severe (A-M-S) progress groups, 2) initial infection status of HCW pool; 3) infectivity levels (*b_0_, b_1_, b_2_*) for (*E, I_1_, I_2_*) stages and susceptibility levels of individual hosts or host-pools; 4) average duration of infective stages for (A-M-S) pathway; 5) daily social mixing patterns between HCWs and infected patients; 6) daily isolation of symptomatic cases and recovery (See SI in details).

### Model calibration

Our model is calibrated to empirical data of a COVID-19 outbreak among HCWs in the department of neurosurgery of Union Hospital, Wuhan, China from January 5th, 2019 to February 4th, 2020 [30]. A Bayesian method is used to calibrate the following important parameters in our IBM: (i) mean infectivity (*b_1_,b_2_*) of symptomatic hosts (*I_1_,I_2_*), (ii) increased susceptibility level (0.5 < *a_S_ <* 1) of the high-risk pool; (iii) fraction *v_A_* of HCWs going through the asymptomatic pathway (*E → R*).

The Bayesian method uses the posterior probability distribution to quantify the uncertainties in these model parameters using the observed data on the daily incidence of symptomatic cases and the daily isolated cases. The prior distributions for all these parameters are taken to be uniform within acceptable ranges. The likelihood for the observed data is assumed as a normal distribution with the center at the predicted values from the IBM. The adaptive Metropolis algorithm [33] is used to sample from the posterior distribution, where the jump size is adaptively chosen based on the sample covariances. The chains are run for 10000 iterations, and after 5000 burin-in every 50th sample is used as the final sample from the posterior distribution. To assess the convergence of the posterior sampling, the Gelman-Rubin statistic [34] is computed for all the parameters. The statistics are found to be very close to 1, the desired value in strong support of convergence. The calibration was implemented using R statistical software.

### Wuhan Hospital outbreak

On December 26th, 2019, a patient later diagnosed with COVID-19 was admitted in the department of neurosurgery of Union Hospital, Wuhan, China. No PPE was used by HCWs at that time. By January 8th, HCWs started to show COVID-like symptoms (headache, cough, sore throat), and screening and isolation were initiated among HCWs. From January 19th, patient’s admission was stopped in the department, and the hospitalized patient pool was gradually reduced from 200 to 20 by the beginning of February. Over the period from January 5th to February 4th, 92 out of the 171 HCWs of the department were suspected or confirmed COVID-19 cases and isolated. New patients were only admitted in early March 2020 when the pandemic was declared under control in Wuhan.

### Intervention strategies

We consider three types of interventions: (i) social distancing (reduced contact rates) among HCWs and individual protection (facial masks), (ii) enhanced screening and isolation of infected HCWs, (iii) patient-pool control (pool size and infection level), and individual HCW protection via PPE.

For our baseline case, we assumed 50% and 100% isolation (quarantine) fractions of symptomatic cases (*I_1_; I_2_*), respectively, and fixed infection level of the patient pool (see Table S1). To account for model uncertainties, we run each control simulations for 100 posterior parameter samples and 5 stochastic model realizations for each sample (500 histories altogether), over a six-month period.

For social distancing, we considered 50% and 75% contact-rate reduction relative to their baseline values. The effect of face mask on inter-staff or staff-patient mixing, was simulated by reduced susceptibility of individual HCWs, with several values of mask efficacy [9]. Screening and isolation fractions (*f_i_*) of HCWs were based on limited test sensitivity, combined with non-COVID symptoms. An increase in targeted isolation assumes more intensive screening or test sensitivity. We also studied the effect of isolating pre-symptomatic/ asymptomatic cases (*E* –pool). This task is more challenging, as PCR tests have lower sensitivity for such hosts [32], so to identify a suitable *E* –fraction would require intensive mass screening or contact tracing.

For quantitative assessment of control interventions and their impact, we use two measures: (1) outbreak size = infection turnover (by the end of outbreak); (2) workday loss estimated from the quarantine pool over the outbreak duration. The latter gives a simple economic measure of outbreak impact and putative interventions. In each control experiment, we compare the ratio of two outputs (outbreak size and workday loss) to their baseline values, and record these relative values and their distribution.

Another important factor in the hospital setting is the in-patient pool. In our case (a non-COVID unit in Wuhan), it varied from the full capacity to zero. The key inputs of the patient pool included (i) infected prevalence, (ii) mean patient infectivity to HCWs. The former is controlled by patient admission and screening/isolation procedures; the latter can be modulated by using PPE. We also explored the effect of different timing of PPE implementation and its efficacy.

## Results

### Model calibration

The predictions from the calibrated IBM were very close to the observed data on daily symptomatic and quarantine cases (see Figure 2). The fraction of asymptomatic disease-progress pool, *v_A_*, was estimated at 0.308 (90% Credible Interval (CrI): 0.163 – 0.395). So a sizable part of transmission was carried over by undetected cases (*E* –pool). Susceptibility level of the high-risk pool was estimated at *a_S_ =* 0.76 (90% CrI: 0.582 – 0.965). We attributed a higher susceptibility level to work-stress, and our results gave a quantitative measure to this increase at 52% (90% CrI: 16.4% – 93.0%) above the normal level. The infectivity levels of pre-symptomatic and symptomatic infections were estimated to be 0.124 (90% CrI: 0.111 – 0.144) and 0.225 (90% CrI: 0.202 – 0.262). See SI Figure. S3 for the prior and posterior probability distributions.

**Figure 2:**
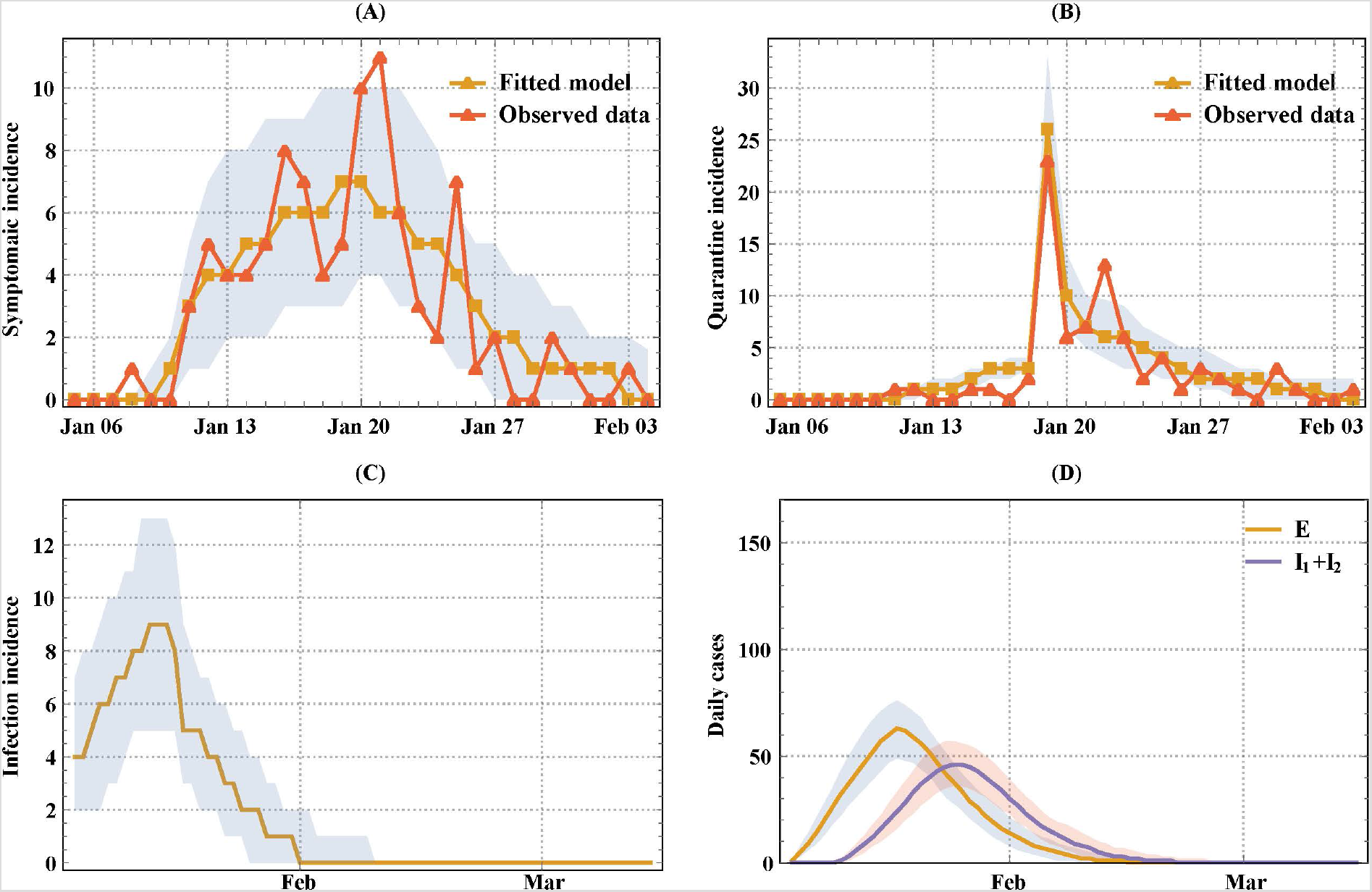
Model calibration and prediction. Panel A shows the observed and fitted daily incidence of symptomatic cases (*E → I_t_*). Panel B shows the observed and fitted daily quarantine cases among HCWs. Panel C shows the predicted infection incidence (*S → E*) from the calibrated model. Panel D shows the corresponding predicted daily pre-symptomatic/ asymptomatic cases, *E*, and symptomatic cases, *I_1_ + I_2_*, respectively. The grey shaded regions are 90% credible intervals.

### Interventions

The baseline scenario showed that almost all HCWs get infected, resulting in significant workday loss, 1050 (90% CrI: 913–1282) over the six-month period (Table 1 and SI Figure. S4). The impact of implementing social distancing through reduction of contact rates alone and wearing facemasks alone, from the start of the outbreak, was evaluated (Table 1). The reduction of contact rates alone has a marginal effect on mitigating the outbreak in the long run. The 50% - drop of contact rates leads to about 4%-6% reduction of the outbreak size and workday loss, while 75% - drop leads to a 15%-17% reduction, relative to baseline values. The efficacy of facemasks is uncertain, and we explored several values (50%, 67%, 75%, 85%, 95%), based on the previous studies [9]. We have shown that wearing facemasks had a higher impact on mitigating the outbreak, than social distancing (reduced contact rates). At 95% efficacy, we could achieve 80% (90% CrI: 73.1% – 85.7%) reduction of outbreak size, and 87% (CrI: 80.0% – 92.5%) of workday loss, compared to the baseline.

**Table 1:** Effects of implementing social distancing through reduction of contacts alone and wearing face masks alone, from the start of the outbreak. We simulated a six-month intervention-regimen for the calibrated model. The progress was measured in terms of outbreak size, workday loss, and cumulative quarantine incidence. Reasonable (50% and 75%) reduction of contact rates and levels of efficacy of facial masks (50%, 67%, 75%, 85%, 95%) were chosen. The results shown are predicted median (90% Credible Interval).

| Progress Measure | Intervention |  |  |  |  |  |  |  |
| --- | --- | --- | --- | --- | --- | --- | --- | --- |
|  | Baseline Results (no intervention) | Reduction of contact rates |  | Efficacy of facial masks |  |  |  |  |
|  |  | 50% | 75% | 50% | 67% | 75% | 85% | 95% |
| Outbreak size | 170 | 164 | 146 | 159 | 141 | 124 | 90 | 35 |
|  | (168-171) | (159-169) | (135-156) | (150-165) | (128-151) | (108-137) | (75-104) | (24-46) |
| Workday loss | 1050<br>(913-1282) | 992<br>(855-1202) | 853<br>(721-1044) | 950<br>(817-1145) | 819<br>(678-997) | 698<br>(562-876) | 469<br>(342-605) | 142<br>(77-212) |
| Cumulative<br>quarantine<br>incidence | 112<br>(98-136) | 107<br>(93-130) | 93<br>(80-113) | 103<br>(89-123) | 90<br>(76-108) | 78<br>(63-96) | 53<br>(40-68) | 16<br>(9-24) |

Figure 3 illustrates the combined effect of facemask and social contact. We used the same values of facemask efficacy and contact rates as Table 1. For each value of facemask efficacy, we observed a consistent reduction of the outbreak size with reduced contact rates. It varied from 13% – 34% drop for low-efficacy facemask (50% protection), to 30% – 60% drop for high-efficacy facemask (95% protection). We observed a similar percentage reduction for the workday loss. So the impact of reduction of contact rates was much greater under the higher efficacy of facemasks.

**Figure 3.**
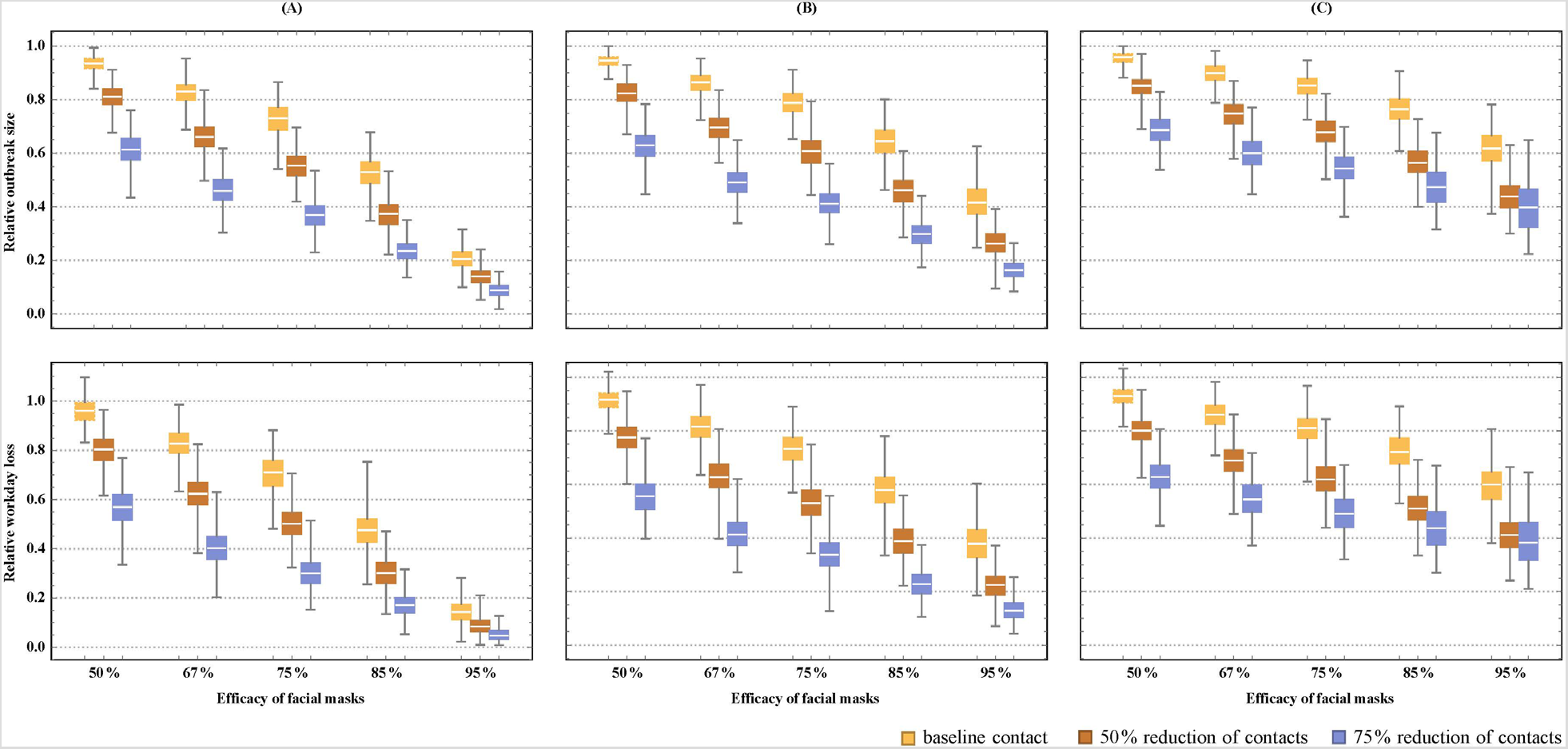
The combined effect of facial masks and social distancing (reduced contact rate). Three levels of social distancing were considered (normal, reduced by 50%, reduced by 75%). For face-mask efficacy, we considered five putative values (50%, 67%, 75%, 85%, 95%). The efficacy is measured via reduced host susceptibility per contact (a → *0 • 5 * a; a • 0 * 33 * a…*). We also considered different timing of preventive measures: (1) start of the outbreak (Panel A); (2) after the first identified HCW case (Panel B); (3) after 10% of HCW-staff got infected (Panel C). In each case, we estimated the posterior distribution of the relative outbreak size, and the workday loss over baseline values.

We also explored the effect of timing of intervention by the following three scenarios: (1) at the beginning (Figure. 3A); (2) after the first identified case (Figure. 3B); (3) after 10% of HCWs have been identified as infected (Figure. 3C). Early interventions have made marked improvement under different types of facemasks and contact rates. For instance, if control interventions (adoption of high-efficacy facemasks and reduced contact rates) were implemented at the start of the outbreak, we observed 80% – 90% reduction of the outbreak size (a near-complete control). A later implementation (e.g. after the first identified case), gave 60% – 85% reduction. If the timing was delayed to e.g. 10% identified cases, these numbers dropped to 40% – 60%. All intermediate cases were shown in Figure. 3.

We next looked at the effect of HCWs screening and isolation via two scenarios. The first scenario considered symptomatic cases only, by changing quarantine fraction (*f*_1_) of *I*_1_, from its baseline value (50%) to *60% → 100%;* quarantine fraction (*f*_2_) of *I*_2_ was fixed at 100%. Figure. 4A shows increased symptomatic isolation had only a marginal effect on the outbreak size, while raising workday loss. A clue to low efficacy of symptomatic screening lies in (i) the role of pre-symptomatic/ asymptomatic (E) pool in transmission, (ii) contribution of the patient source. To test (i) we extended our quarantine strategy to pre-symptomatic/ asymptomatic cases (*E*). Of course, such an extension requires intensive screening of the work pool. Under random selection, isolating *f* –fraction of *E*, would require much more than *f* –fraction of HCWs tested. For numeric simulations, we fixed (*f*_1_, *f*_2_) at (90%, 100%), and varied *E* –fraction from 10% to 60%. We still found the effect of such a strategy was limited, it often prolongs the outbreak duration without affecting its size. Besides, such a strategy can incur an economic burden by increased workday loss, though the effect is subtler, as increased quarantine rate can slower transmission rate, hence fewer hosts would be infected and need isolation. More significant progress was achieved by controlling the patient source, via reduced patient prevalence (screening), or reduced infectivity (PPE) (see Figure. 4A–B).

**Figure 4.**
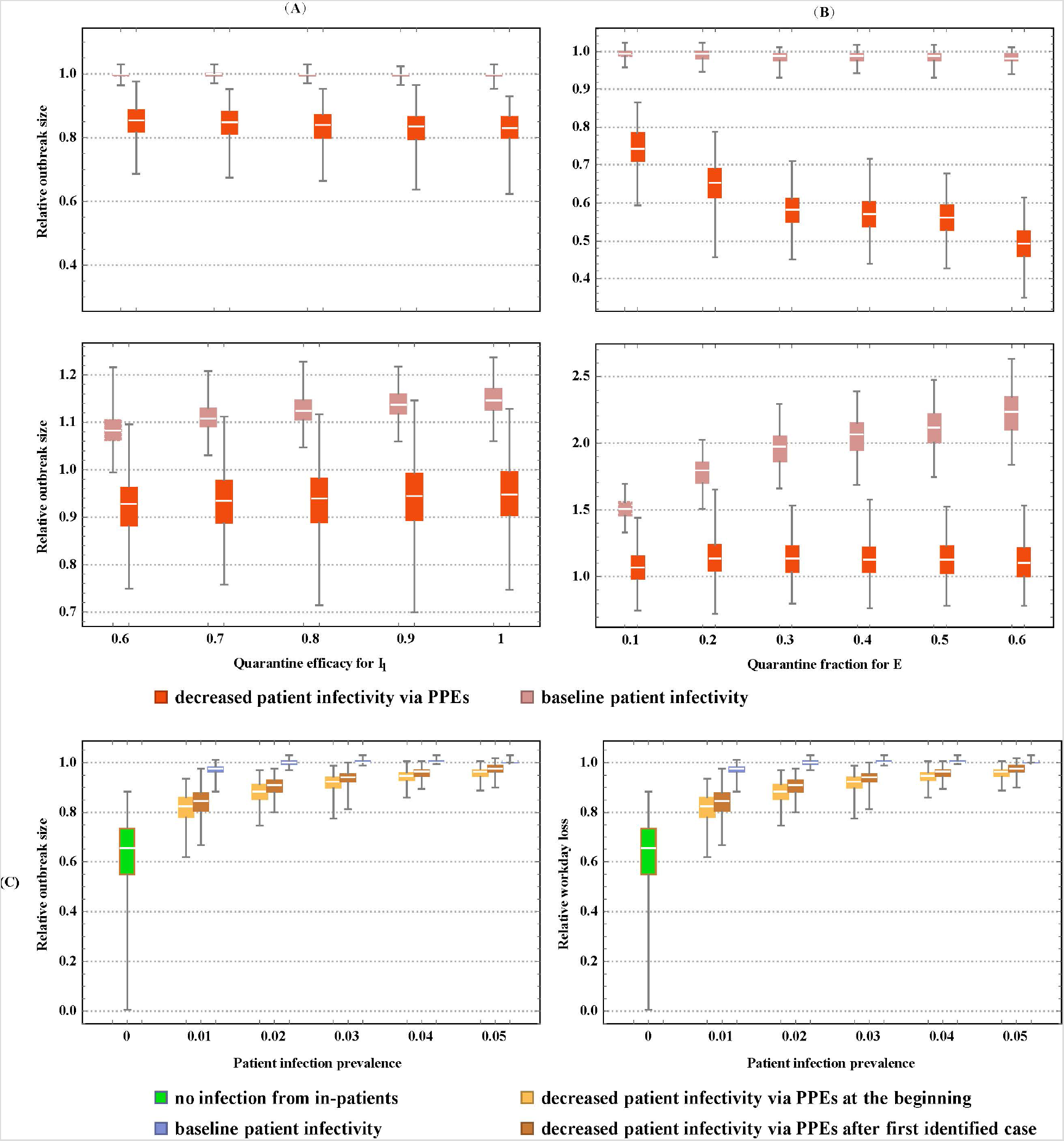
The effect of the quarantine and patient sources on relative outbreak size and workday loss. The patient sources (infection prevalence and infectivity) were controlled via screening/isolation and the use of PPE by HCWs. We considered two quarantine strategies for HCWs: symptomatic cases only (column A), adding pre-symptomatic/ asymptomatic cases (column B). We used the following marking: (pink) baseline patient infection level, (red) reduced patient infection by 80% via PPE use by HCWs. In column A, the quarantine fraction of moderate/severe cases (*l_2_*) was fixed at 100%, and the quarantine fraction of mild cases (*1-^*) was varied from 60% to 100%. In column B, we fixed symptomatic (*1^, I_2_*) quarantine fractions at (90%, 100%), and varied the quarantine fraction of pre-symptomatic/ asymptomatic £-pool from 10% to 60%. Panel C shows the effect of patient infection and different timing of PPE use: i) start of the outbreak, ii) after the first identified HCW infection, iii) no PPE use. We considered different levels of prevalence of the infected patient pool: 0%, 1%, 2% (baseline value), 3%, 4%, 5%.

We run several experiments with patient-pool control and PPE use (Figure. 4C). For PPE timing, we made three choices: (i) the start of an outbreak, (ii) after the first identified HCW-case, (iii) no PPE use. We assumed PPE provides 80% protection (via the reduced probability of transmission from an infected patient). We also varied the infected prevalence level of the patient pool, from 0% to 5% (baseline case was 2%). We found the control of patient infection (via e.g. PPE, screening and isolation, particularly for new patients) can reduce outbreak size, even though the bulk of transmission is carried over by inter-staff HCW contacts. We found the combined strategy (enhanced HCW screening/isolation with patient control) could lead to marked improvement both in outbreak size and in workday loss. This effect, however, is not observed for quarantine alone, under persistent patient source.

Overall, we saw high-efficacy facemasks could provide the most effective control tool for reducing COVID-19 transmission in HCW staff (Figure. 3).

## Discussion

With the spread of COVID-19 in the world, the development of strategies for mitigating its severity is a top public health priority. Large-scale population-level models of SARS-CoV-2 transmission can give some qualitative answers for outbreak control on regional/country scales [35], however, few studies have looked at the effects of interventions in a local community setting, such as hospital, workplace, and school.

Using a novel individual-based modelling approach, we explored different scenarios for COVID-19 transmission and control in a non-COVID hospital unit. Our IBM methodology employed conventional SEIR disease-stages with graded infectivity, extended to heterogeneous host makeup, which includes multiple disease pathways, varying individual susceptibility, and behavioral patterns. These factors can be affected by work stress, health status, use of face masks /PPE, and social interactions. Social mixing was implemented in our model via inter-HCW contact pools and HCW-patient interactions. Control interventions included test diagnostics and isolation of established cases. Detailed data on the COVID-19 outbreak in the department of neurosurgery of Union Hospital in Wuhan (China) [30] was used to calibrate the essential model parameters. It included the onset of symptoms, diagnosis and isolation, and patient pool control, over the period January 5th, through February 4th, 2020.

We allowed different disease-progress pathways: severe, mild, and asymptomatic (S-M-A), so the HCW-pool was partitioned into (S-M-A) progress groups. One of the key uncertain parameters was the asymptomatic-fraction, estimated at 31%, a relatively high proportion of undetectable infections. Another uncertain input was individual susceptibility, which could be affected by health status or work stress. Based on data analysis [30], we divided the work pool into a normal group (60% of HCWs), and a high-risk group (40% of HCWs). We estimated the high-risk susceptibility level relative to normal susceptibility and found work-related stress could increase the risk of COVID-19 infection by up to 52%.

The calibrated model was used to simulate a range of intervention scenarios, aimed at mitigating the outbreak and examining its impact on the work pool. The baseline case, without interventions, gave a large outbreak size, whereby almost all HCWs were infected over two months. It also incurred a significant workday loss for the unit. Our results support early modeling findings of large-scale populations, and subsequent empirical observations, that in the absence of control measures, a COVID-19 epidemic could quickly overwhelm a region [12]. High-efficacy facemasks were shown to be most effective for reducing infection cases and workday loss. The impact of social distancing through the reduction of contact rates alone had an only marginal effect on mitigating the outbreak in the long run. Reducing social contact rates to 50% (or 70%) resulted in a 4%-6% (or 15%-17%) drop in the outbreak size, and a similar drop in the workday loss, compared to the baseline case. However, the impact of reduction of contact rates was much greater under the higher efficacy of facemasks.

Implementing the quarantine policy (HCW screening and isolation) alone, even when all symptomatic cases are included, would typically prolong the outbreak duration, but had a marginal effect on its size, particularly under the existence of external (patient) source pressure. Our results indicated that the low efficiency of symptomatic quarantine was due to a large share of transmission being carried by pre-symptomatic/ asymptomatic (*E*) individuals [36], and to the patient source. Hence, a quarantine policy for HCWs should be augmented with other interventions to achieve a significant reduction. Efficient control of the patient source (via the use of PPE, their screening and isolation, and/or admission) is one key to mitigating the HCW outbreak. The effectiveness of all these interventions was shown to increase with their early implementation.

To our knowledge, this study is the first of its kind to provide quantitative modelled assessment and projections for COVID-19 transmission in hospital settings. However, the IBM methodology developed here, has a far broader scope, beyond healthcare facilities. Indeed, with proper adjustment, it could be applied to many other local communities (workplaces, schools, city neighborhoods, et al). The key feature of such IBM is a fine-scale resolution of community makeup, social interactions, and disease pathways. Such information is essential for risk assessment and the development of efficient control/ intervention strategies on a local scale.

The current model setup is subject to some limitations. First, it was designed for a single hospital unit and simplified treatment of the patient pool, as the target group in our study was HCW-pool. More realistic local communities could combine multiple units (e.g. large hospital), with refined population structure (e.g. patients, visitors, staff), and more complex interactions (e.g. ‘random’ and ‘scheduled’ contact pools). Empirical data on these interactions will be required to adequately parameterize such models. Second, although we have made an effort to characterize the SARS-CoV-2 transmission in a hospital setting, some parameters used in our setup were drawn from general information sources, such as fractions of symptomatic mild and severe cases [37], disease stages and durations [38], and associated infectivity levels [39], which may be adjusted in the future work.

## Conclusion

Overall, our analysis shows that a COVID-19 outbreak among HCWs in a non-COVID-19 hospital unit can be efficiently controlled /mitigated by non-pharmaceutical means. The most crucial factor of success is high-efficacy facemasks for HCW contacts. It can be further augmented by social distancing, screening/isolation, and patient source control.

## Data Availability

The computer codes, including the aggregated data, implemented in the Wolfram Mathematica platform are available in Githubs https://github.com/qimin-h/COVID-19-huang-et-al.-. All other materials are available from the corresponding authors on reasonable request.

## List of abbreviations

WHO: World Health Organization
HCWs: healthcare workers
PPE: personal protective equipment
CrI: Credible Interval

## Declarations

### Ethics approval and consent to participate

The study protocol was approved by the institutional ethics board of Union Hospital, Tongji Medical College, Huazhong University of Science and Technology, Wuhan, China (No. 20200029). Written informed consent was required before the data collecting, and participants were informed that they could refuse to answer any question. The questionnaire did not ask about infection status, and no biological samples were collected.

### Consent for publication

Not applicable.

### Authors’ Contributions

QH, AM, and XJ contributed equally and shared the first authorship. QH, DG, AM, and MN designed research, did model development, calibrations, and simulations, XJ, HZ, FF, PF, and WX collected and provided hospital data, QH wrote the first draft, MN, DG, AM, and MAH made critical revision of the manuscript. All the authors contributed to the interpretation of the study results, read, comment, and approved the final version.

## Acknowledgments

The authors would like to appreciate all healthcare workers in this study. XJ and HZ had full access to all the data in the study and took responsibility for the integrity of the data.

## Supplementary information

### Setup and parameters of the individual-base model

The individual-based model we developed features a heterogeneous host population with distinct infective stages and transitions, different patterns of disease-progression (asymptomatic, mild, severe), individual risk factors (health status, work stress, et al), individual behavior including social mixing patterns among HCWs, use of PPE/ facial masks, and HCW-patient interactions.

#### The key inputs in IBM are described below

1. Host community consists of three disease progress groups, asymptomatic, mild, severe (A-M-S), i.e., population fractions (*v_A_, v_M_, v_s_*) that undergo three pathways of Figure 1: (A) asymptomatic (*E → R*); (M) mild symptomatic (*E →I*_1_ → *R*); (S) severe symptomatic (*E → I*_1_ → *I*_2_ → *R*). Based on the WHO report [1], we assumed that for symptomatic cases, 80% are mild symptomatic, i.e., *v_M_* = 0.8 * (1 - *v_A_*), *v_s_* = 0.2 * (1 - *v_A_*), Fraction *v_A_* was estimated at 0.3080 (90% Credible Interval (CrI): 0.1628 – 0.3948) using the Bayesian method, consequently, we get (*v_M_, v_s_*) equal to (0.5536; 0.1384);
2. Initial infection state. We assume that only one HCW was in the latent E stage at the beginning of the simulation, while the rest 170 HCWs were susceptible;
3. Stage-specific infectivity levels (*b*_0_, *b*_1_, *b*_2_) for (*E, I*_1_, *I*_2_). We assume *b*_1_ = *b*_2_, for symptomatic groups (*1^ I*_2_), while asymptomatic infectivity *b*_0_ = 0.55 * *b*_1_ [2]. Then we calibrated *b*_1_ to be 0.2245 (90% CrI: 0.202 – 0.262) using the Bayesian method, consequently, *b*_0_ = 0.124 (90% CrI: 0.111 – 0.144);
4. Individual susceptibility level (*a*). In general, *a* may depend on host health /immune status, individual behavior e.g. use of facial masks or PPE, and environmental conditions, *a* = 0 (fully protected), *a* = 1 (fully susceptible). Work stress is another factor that can affect susceptibility. According to [3] around 40% HCWs claimed stress during the outbreak. Hence, we allowed 60% HCWs to have a baseline susceptibility, *a_N_* = 0.5, while high-risk(stressed) pool, 40% of HCWs, with susceptibility level 0.5 < *a_S_* < 1 (to be calibrated by the Bayesian method).
5. The average duration of infective stages for each pathway. According to the published studies [1, 4–8], we fixed the average duration *L_ER_* of asymptomatic path (*E → R*) to be 10 days; the average durations (*L*_*EI*_1__, *L*_*I*_1_*R*_) of mild symptomatic path (*E → I*_1_ → *R*) to be (5;9) days; and the average durations (*L*_*EI*_1__, *L*_*I*_l_*I*_2__, *L*_*I*_2_*R*_) of severe symptomatic path (*E → I*_1_ → *I*_2_ → *R*) to be (5; 3; 14) days.
6. Social mixing patterns between HCW, and external patient sources; Intra-HCW contacts were simulated as daily aggregation in small groups of 2,3, or 4 hosts, randomly drawn from the current working HCW pool. The basic case (full HCW working pool) assumes 60 pair-contacts, 30 triple-contacts, 8 quadruple-contacts per day. It approximately gives a 2.2 contact rate per HCW-host per day from hospital data (Figure S2) [3]. Larger host aggregations are possible, but we ignore them here. Besides internal mixing, each HCW-host visited patients at a prescribed rate = 20/per day. Unlike HCW staff, the patients were not individualized, but a random patient cluster (determined by mean visitation number per day) was drawn from the total patient pool (200) with a prescribed infected fraction (0.02), using a hypergeometric distribution. According to hospital data, no new patients were admitted after January 19th, and their pool was discharged after January 19th, at a rate of 5% /day, and HCWs will wear PPEs in face of patients after Jan 19th, so we assume patient infectivity, *b_PPE_*, is decreased to be 0.02, i.e., the efficacy of PPE is about decreasing risk of infection by 80%, when we calibrated our IBM.
7. Daily isolation of symptomatic cases and recovery. We assumed that HCWs expressing symptoms are tested, and prescribed fractions (*f*_1_; *f*_2_) of (*I*_1_; *I*_2_) put in isolation. The assumed quarantine fractions (0 < *f_t_* < 1) combine limited test sensitivity, and overlapping ‘COVID-like’ symptoms, expressed by other (non-COVID) hosts. According to data, the Union Hospital in Wuhan has much more strict quarantine policy after January 19th, when they were aware of the seriousness of COVID-19, so we fixed *f*_1_ to be 0 1 and 08 before and after January 19th, respectively, and fixed *f*_2_ to be 0.15 and 0.85 before and after Jan 19th, respectively, when we calibrated our IBM.

**Table S1.** IBM inputs and parameters.

| Parameter | Descriptions | Value | Source |
| --- | --- | --- | --- |
| $(a_N; a_S)$ | Susceptibility of a susceptible individual without stress and with stress | (0.5; 0.7608) | Calibrated |
|  | Percentage of HCWs without and with stress | (0.6,0.4) | Data [3] |
| $(b_0, b_1)$ | Infectivity of an asymptomatic and symptomatic infectious individual, respectively | (0.1235, 0.2245) | Calibrated [2] |
| $(b_0, b_{PPE})$ | Infectivity of patient before January 19th and after January 19th (because of PPE), respectively | (0.1235, 0.02) | Estimated based on hospital policy |
| $(n_2, n_3, n_4)$ | Number of random pair, triple, quadruple contacts per day among HCWs | (60;30;8) | Typical contacts/day |
| $L_{ER}$ | The average duration of asymptomatic ($E \rightarrow R$) | 10 | [1, 4-8] |
| $(L_{EI_1}, L_{I_1R})$ | The average duration of mild symptomatic ($E \rightarrow I_1 \rightarrow R$) | (5;9) | [1, 4-8] |
| $(L_{EI_1}, L_{I_1I_2}, L_{I_2R})$ | The average duration of moderate/severe symptomatic ($E \rightarrow I_1 \rightarrow I_2 \rightarrow R$) | (5;3;14) | [1, 4-8] |
| $q$ | Transition rate of disease progression based on average duration and days | $\frac{(d/L)^m}{1 + (d/L)^m}$ | |
| $f_1$ | Screening/ diagnostic fraction of mild symptomatic infectious population before and after January 19<sup>th</sup>, 2020, respectively; | (0.1;0.8) | Estimated based on hospital policy. |
| $f_2$ | Screening/ diagnostic fraction of severe symptomatic infectious population before and after January 19<sup>th</sup>, 2020. | (0.15;0.85) | Estimated based on hospital policy. |
| $(v_A, v_M, v_S)$ | Fraction of populations goes through three paths of disease progression (1) asymptomatic ($E \rightarrow R$); (2) mild symptomatic ($E \rightarrow I_1 \rightarrow R$); (3) moderate/severe symptomatic ($E \rightarrow I_1 \rightarrow I_2 \rightarrow R$) | (0.3080; 0.5536; 0.1384) | Calibrated [1] |
| **External source from patients** |  |  |  |
|  | average number of visits to patients per HCW per day; the number of infectious patients, the total number of patients | (20;4;200) | Data |
| **Initial symbols** | **Description** | **Values** | **Source** |
| $(S_0, E_0)$ | initial susceptible population; initial exposed population | (170;1) | Data |

**Figure S1.**
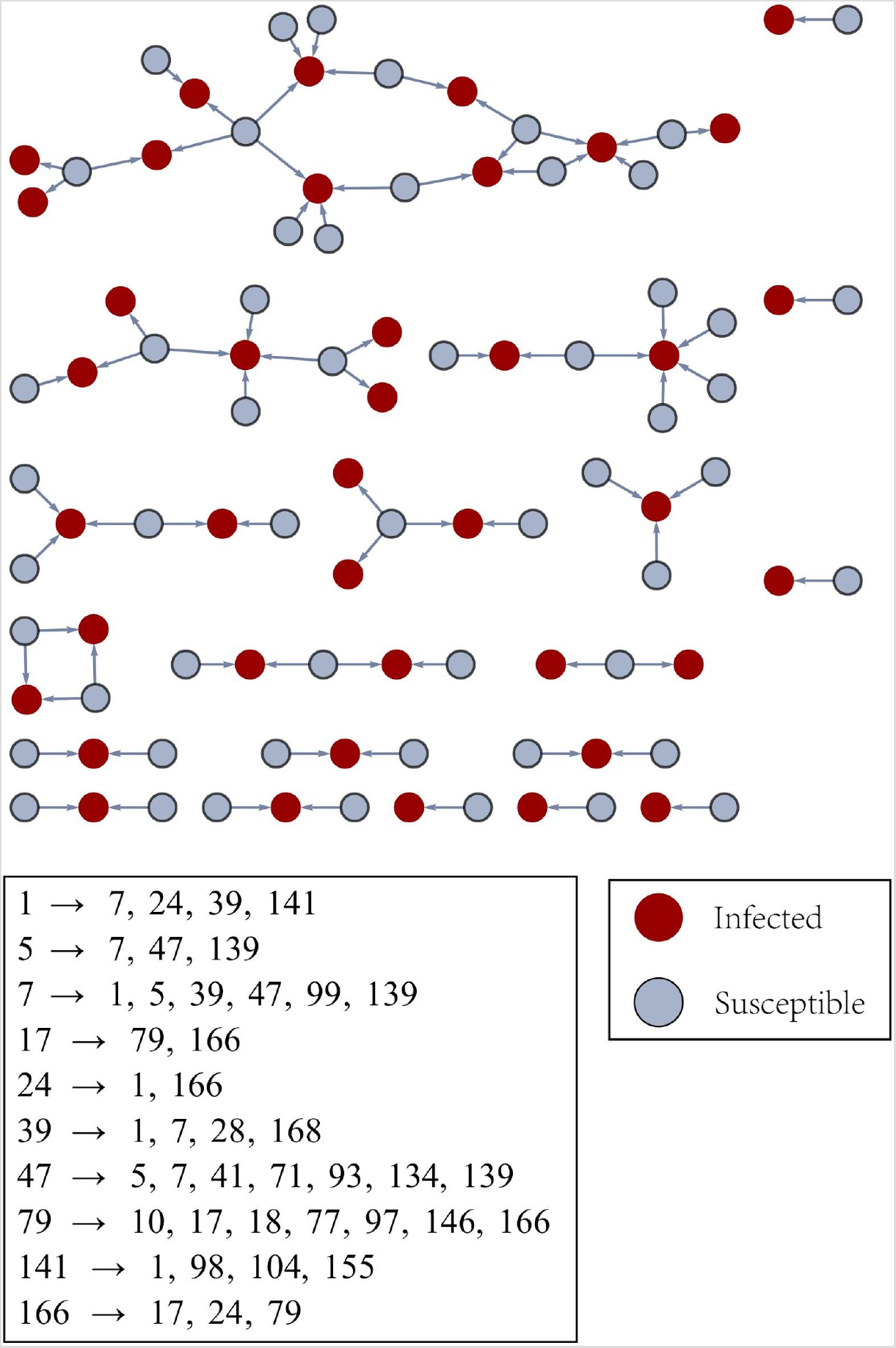
A typical daily snapshot of transmission pathways resulting from random mixing. Table at the bottom shows selected contact pools, and the graph arrows indicate which susceptible hosts (grey) were potentially in contact with infective (*E → I*) hosts (red). In our scheme, a single ‘red’ can infect multiple susceptibles, and a susceptible (grey) can be linked to multiple infectives.

**Figure S2.**
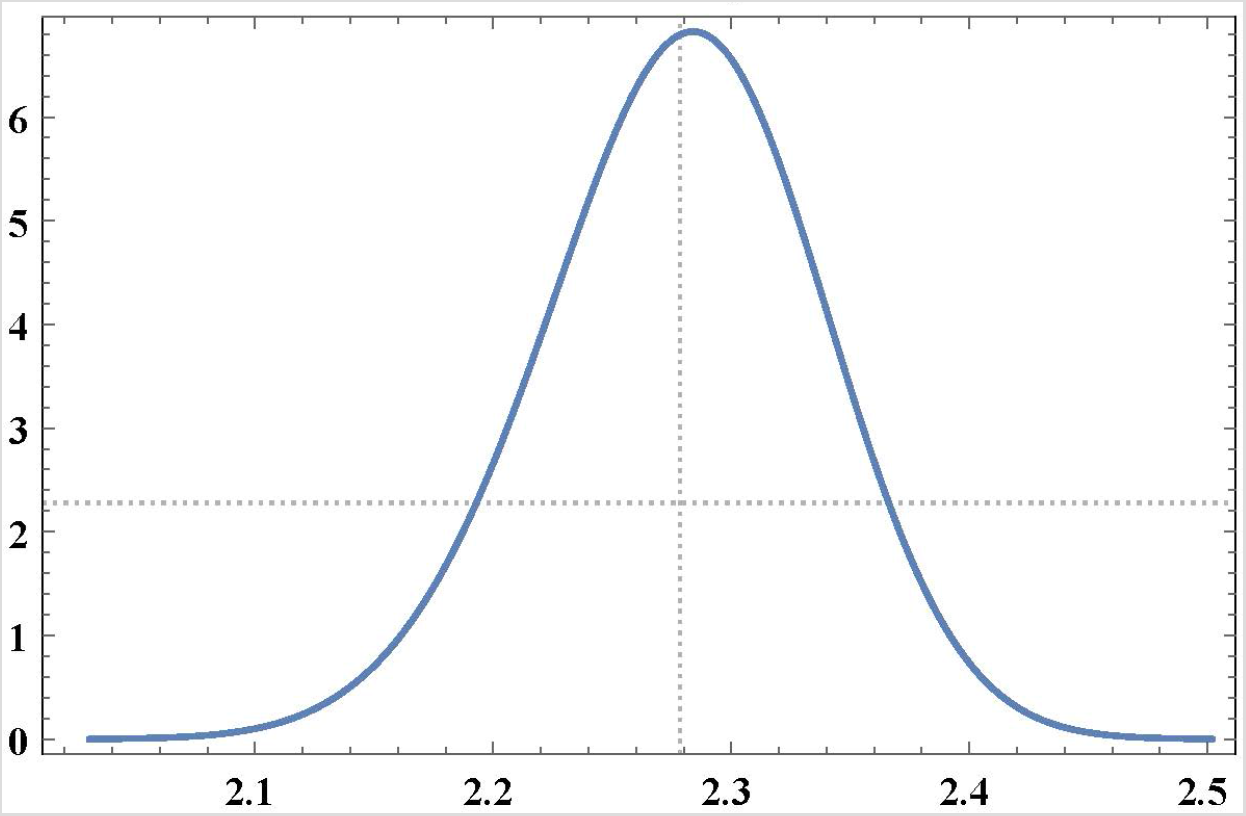
Distribution of baseline contact. The basic case (full HCW working pool) assumes 60 pair-contacts, 30 triple-contacts, 8 quadruple-contacts per day. It approximately gives a 2.2 contact rate per HCW-host per day.

**Figure S3.**
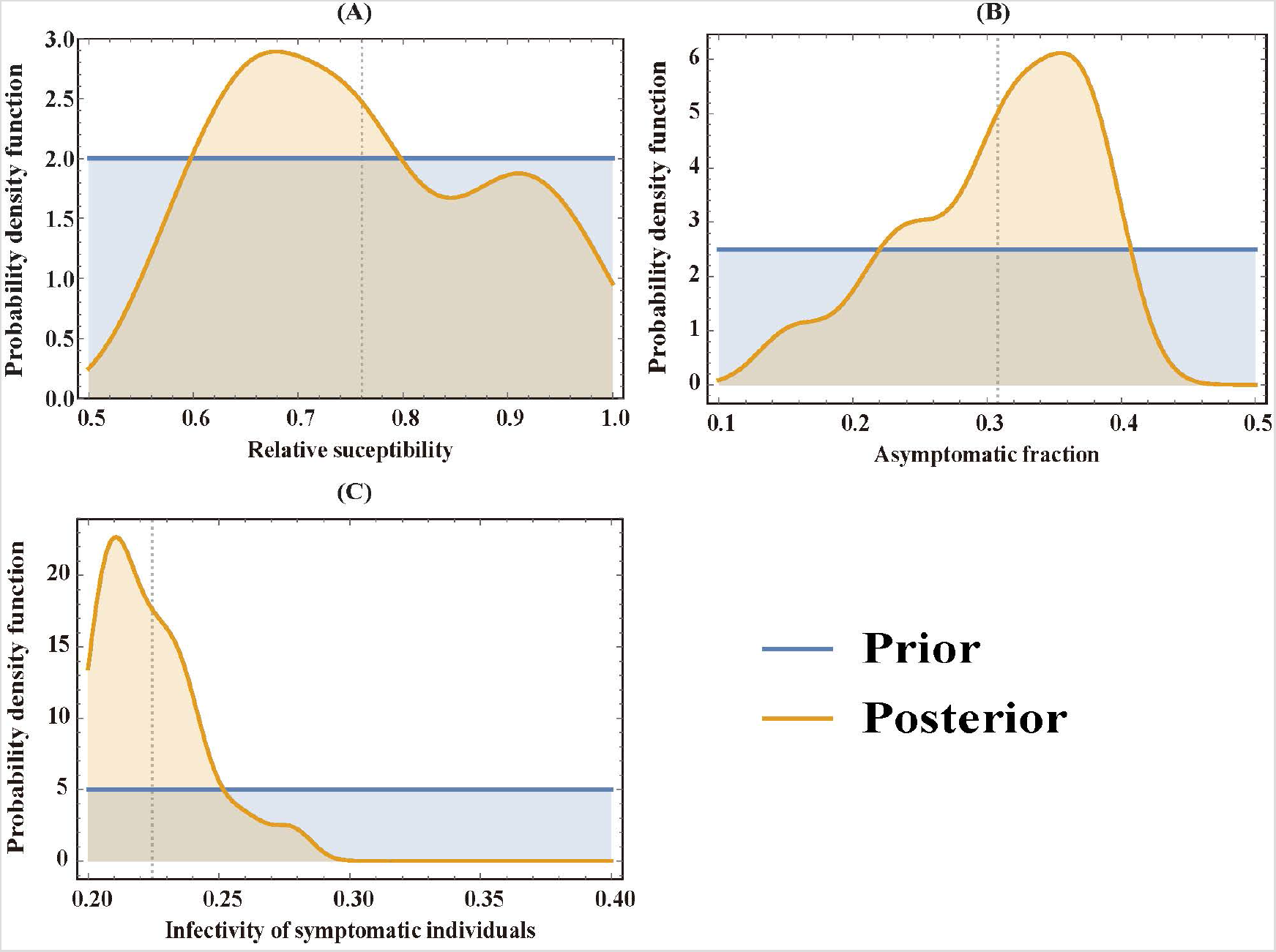
The prior and posterior distribution for key parameters. Panel (A): relative susceptibility, *a_s_*, of the stressed HCWs when compared to a baseline susceptibility of *a_N_* = 0.5; The prior distribution of *a* was uniform in (0.5,1) and the posterior was estimated to be 0.7608 (90% CrI: 0.5822 – 0.9653). Panel (B): asymptomatic fraction, *v_A_*, i.e., the fraction of the HCWs which goes through the asymptomatic pathway (*E → R*). The prior distribution of *v_A_* was uniform in (0.1, 0.5) and the posterior was estimated to be 0 3080 (90% CrI: 0.1628 – 0.3948). Panel (C): infectivity of a symptomatic individual, *b_x_*. The prior distribution of *b_t_* was uniform in (0.2, 0.4) and the posterior was estimated to be 0.2245 (90% CrI: 0.202 – 0.262).

**Figure S4.**
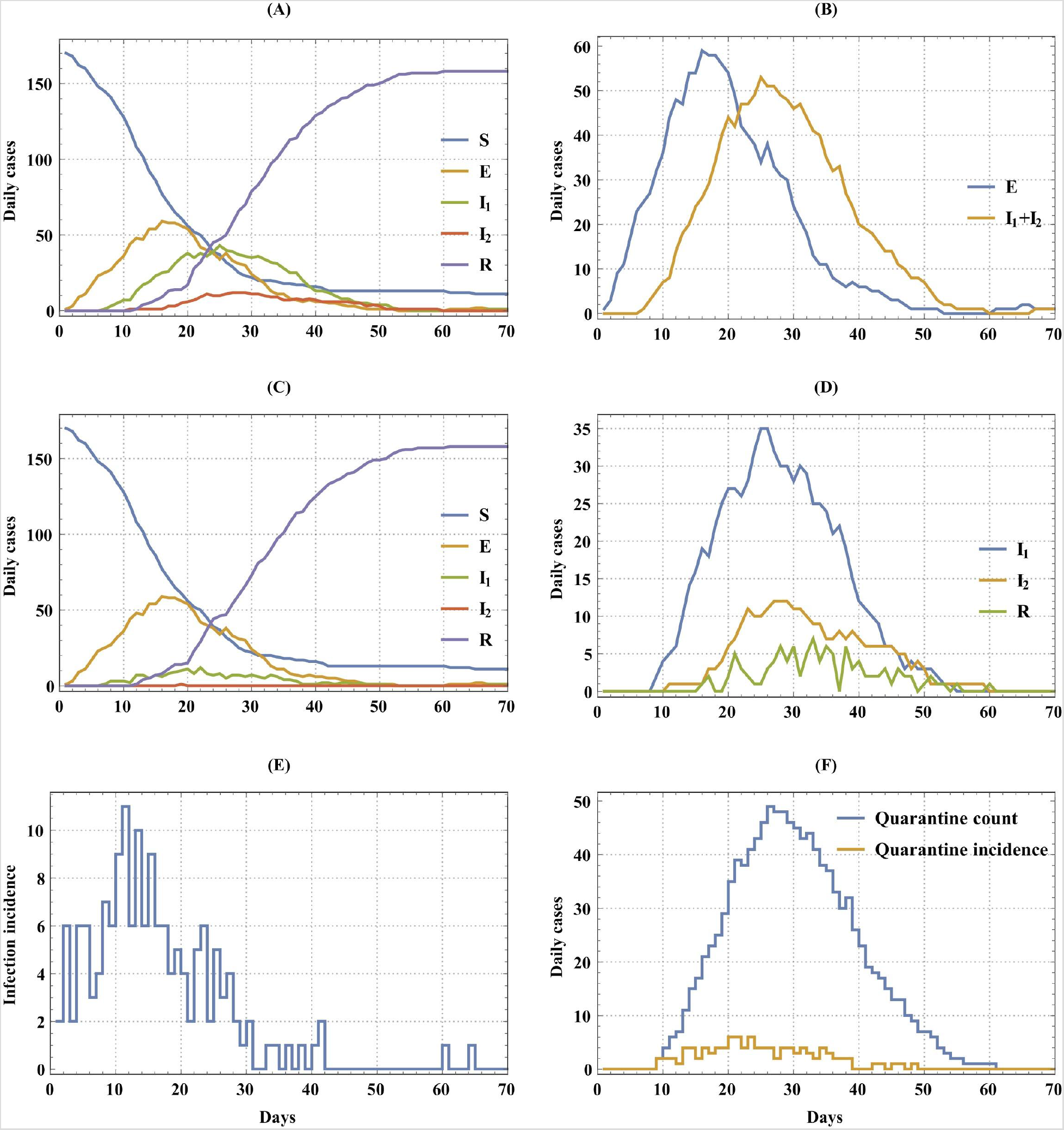
Typical outbreak history over a 70-day period for a baseline case. Panel (A) shows the outbreak history of the combined pool (work pool+quarantine pool); Panel (B) shows the simulation of asymptomatic, *E*, and symptomatic cases, *I_1_ + I*_2_, in the combined pool. Panels (C) and (D) show the history of the work pool and quarantine pool, respectively. Panel (E) shows daily infection incidence (*S → E*). Panel (F) shows daily quarantine incidence (yellow) and quarantine count (blue), respectively.

